# RESiLIENT (Resilience Enhancement with Smartphone in LIving ENvironmenTs) Trial: The Statistical Analysis Plan

**DOI:** 10.1101/2024.03.16.24304261

**Authors:** Hisashi Noma, Toshi A. Furukawa, Aran Tajika, Masatsugu Sakata, Yan Luo, Rie Toyomoto, Masaru Horikoshi, Tatsuo Akechi, Norito Kawakami, Takeo Nakayama, Naoki Kondo, Shingo Fukuma, Helen Christensen, Ronald C. Kessler, Pim Cuijpers, James Wason

## Abstract

This document gives a statistical analysis plan of the acute intervention effects of the RESiLIENT (Resilience Enhancement with Smartphone in LIving ENvironmenTs) trial, designed as a master protocol including four 2×2 factorial trials to elucidate specific efficacies of five cognitive-behavioural therapy (CBT) skills (cognitive restructuring, behavioural activation, problem-solving, assertion training, and behavior therapy for insomnia) using the internet CBT program. This trial was registered within the UMIN Clinical Trials Registry in Japan (UMIN000047124). The study protocol has previously been published (BMJ Open 2023;13:e067850).

## 1. Scope

This document gives a detailed statistical analysis plan of the acute intervention effects of the “RESiLIENT (Resilience Enhancement with Smartphone in LIving ENvironmenTs)” trial and should be read in conjunction with the trial protocol ^1^. Portions of this document are replicated from the study protocol and supplemented with additional detail as appropriate.

## 2. Background of RESiLIENT (Resilience Enhancement with Smartphone in LIving ENvironmenTs) trial

### Objective

Cognitive-behavioural therapy (CBT) is an empirically supported intervention to reduce depression severity and to prevent depression onset ^2-4^. However, it has been traditionally examined as a package of various skills, specific efficacy of which remains largely unexplored. To scale up its application and to combat the rising burden of depression across the globe, we need to assess the specific efficacy of each available CBT skill so that we can inform the most efficacious and efficient intervention package(s). The RESiLIENT (Resilience Enhancement with Smartphone in Living ENvironmenTs) trial ^1^ was designed as a master protocol including four 2×2 factorial trials to elucidate specific efficacies of five CBT skills (cognitive restructuring [CR], behavioural activation [BA], problem-solving [PS], assertion training [AT], and behavior therapy for insomnia [BI]) using the internet CBT (iCBT) program.

### Design

The RESiLIENT trial was designed as a master protocol involving four 2×2 factorial trials examining the efficacy of five smartphone CBT skills. The recruitment started on 1 September 2022 and was completed on 21 February 2024, with the last follow-up of the acute intervention phase planned to complete by 19 April 2024.

This SAP focuses on the effects of iCBT skills up to week 6.

### Study setting

Participants were recruited in the following four fields. The potential participants will access the internet web page for the trial to apply for the study, receive the explanations about the study from the trial coordinators online remotely, and provide their informed consent by electronic signature.

1. Health insurance societies: There are several types of public health insurance schemes together constituting the universal healthcare in Japan. We will collaborate with two large associations, namely, National Federation of Health Insurance Societies covering employees and their families in large corporations (ca 30 million insured), and Japan Health Insurance Association covering employees and their families of small-to-middle-sized corporations (ca 35 million insured).
2. Business companies and corporations
3. Community and local governments
4. Direct-to-consumer advertisements

### Inclusion criteria

1. People of any sex, aged 18 years or older at the time of providing informed consent.
2. In possession of their own smartphone (either iOS or Android).
3. Written informed consent for participation in the trial.
4. Completion of all the baseline questionnaires within 1 week after providing informed consent.
5. Screening PHQ-9 total scores of (1) between 5 and 9, inclusive, or (2) between 10 and 14, inclusive, but not scoring 2 or 3 on its item 9 (suicidality).

### Exclusion criteria

1. Cannot read or write Japanese.
2. Receiving treatment from mental health professionals at the time of screening.

### Interventions

The participants have been randomised equally to one of the following nine intervention arms or three control arms, stratified by employment status (yes or no) using a computer-generated permuted block randomization table implemented in the app.

The intervention arms include:

1. BA
2. CR
3. PS
4. AT
5. BI
6. BA+CR
7. BA+PS
8. BA+AT
9. BA+BI

BA consists of psychoeducation about pleasurable activities according to the principle of “outside-in” under the catchphrase ‘When your body moves, so does your mind’. It provides a worksheet of a personal experiment to test out a new activity and also a gamified ‘action marathon’ to promote such personal experiments.

CR consists of psychoeducation of the cognitive–behavioural model and cognitive restructuring. The participant learns how to monitor their reactions to situations in terms of feelings, thoughts, body reactions and behaviours by filling in mind maps. The participant uses these mind maps to apply cognitive restructuring and find alternative thoughts. In order to help the participant broaden their thoughts, CR provides three tools, each of which guides them to alternative thoughts through interactions with the app characters. PS teaches the participants how to break down the issue at hand, to specify a concrete and achievable objective for it, to brainstorm possible solutions, to compare their advantages and disadvantages, and finally to choose the most desirable action and act on it. A worksheet to guide the participants through this process is provided.

AT consists of psychoeducation of assertive communication in contrast to aggressive or passive communication. The participant learns how to express their true feelings and wishes without hurting others or sacrificing themselves. They fill in worksheets to construct appropriate lines in response to their own real-life interactions.

BI teaches the mechanisms of healthy sleep, invites the participant to keep daily sleep records, based on which the participant will start applying sleep restriction and stimulus control techniques, the two proven behavioural skills to increase sleep efficiency.

Each component consists of seven to nine chapters, each divided into two to four lessons as appropriate, as well as worksheets to practice the skill. The participants are expected to complete one chapter per week and can only move on to the next chapter after a week has passed since they started the previous chapter and after they have completed one worksheet. The participants receive weekly encouragement emails, templated but tailored in accordance with each participant’s progress by the trial coordinators in the management team (all the coordinators do not have a CBT background and are forbidden to provide any therapeutic contents) and are prompted to fill in PHQ-9 every week.

### Controls

In psychotherapy trials, there is no gold standard control condition like the pill placebo in pharmacotherapy trials. Psychotherapy controls must be designed in accordance with the clinical questions of the study as well as the participants’ needs and the available resources, and may produce different effect size estimates (ref). In the RESiLIENT trial, we therefore will use three different control conditions with different levels of stringency.

1. Weekly self-checks
2. Health information
3. Delayed intervention

In the weekly self-check arm, the participants will receive weekly encouragement emails and monitor their moods through the weekly PHQ-9 measurements up to week 6 (as in the intervention arms) and then the monthly PHQ-9 measurements thereafter up to week 50. This arm is intended as an attention control to match the attention provided through encouragement emails and self-checks but lacking in active interventions.

In the health information arm, the participants will receive the URLs of websites containing tips for healthy life (physical activities, nutrition and oral health, none of which focuses on mental health) for the initial 3 weeks and will be asked to answer quizzes for comprehension. They will be asked to fill in self-reports at weeks 3 and 6 (without encouragement emails), and then follow-up evaluations thereafter up to week 50. This arm is intended as a placebo intervention.

In the delayed treatment arm, the participants will be placed on the wait list and be asked to fill in self-reports at weeks 3 and 6. They will then be randomised to 1 of the 11 arms (#1 through #11) at week 6, if they so wish then, and will follow respective programs thereafter.

### Concomitant interventions

All the participants, in the active and control arms, are free to seek whatever mental health interventions they wish through the 50 weeks. The received professional interventions, either pharmacotherapy or psychotherapy, will be monitored and recorded at weeks 6 and 50.

### Primary outcome

The primary outcome is the change in the PHQ-9 from baseline to week 6.

### Secondary outcome

The secondary outcomes include:

1. Changes in PHQ-9 from baseline to weeks 1, 2, 3, 4, and 5.
2. Changes in the Generalized Anxiety Disorder-7 (GAD-7) from baseline to weeks 3 and 6.
3. Changes in the Insomnia Severity Index (ISI) from baseline to weeks 3 and 6.
4. Changes in the Short Warwick Edinburgh Mental Well-Being Scale (SWEMWBS) from baseline to weeks 3 and 6. Anxiety usually coexists with depression and we aim to examine what effects the intervention components, while mainly targeting depression, may have on anxiety. We also aim to monitor broad psychopathology of common mental disorders encompassing depression, anxiety and insomnia as well as positive mental health which are deteriorated in common mental disorders.
5. Changes in the CBT Skills Scale from baseline to week 6.
6. Changes in the Work and Social Adjustment Scale (WSAS), the Utrecht Work Engagement Scale (UWES), the Health and Work Performance Questionnaire (HPQ) and the EuroQOL-5D-5L (EQ-5D-5L) from baseline to week 6.
7. The Client Satisfaction Questionnaire-3 (CSQ-3), adherence to smartphone CBT, co-interventions, safety information at week 6.

## 3. General Considerations

### 3.1 Software

We will use Statistical Analysis System (SAS) Ver. 9.4. (SAS Institute, Inc., Cary, NC) and R Ver. 4.3.2 (R Foundation for Statistical Computing, Vienna, Austria) for all statistical analyses.

### 3.2 Significance Level and Reliability Coefficient

Unless otherwise specified, estimates of treatment effects will be presented with 95% confidence intervals. P-values will be 2-tailed. A P-value <0.05 will be considered statistically significant. No correction for multiple tests will be made.

### 3.3 Numerical Display of Results

As a general rule, the analysis results will be indicated by the number of digits shown below. The data will be rounded to the nearest whole number.

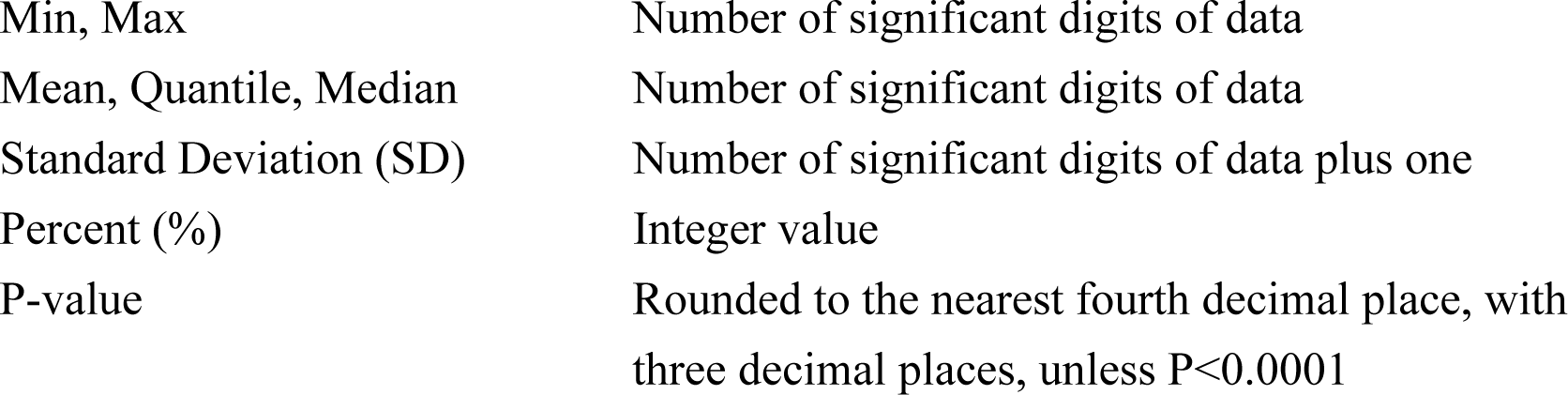

## 4. Data Handling and Analysis Sets

### 4.1 Handling of Data

The handling of individual cases and data will follow the results of examinations by the data safety and monitoring committee.

### 4.2 Analysis Sets

The analysis will follow the intention-to-treat principle and include all the participants randomized.

## 5. Descriptive Analyses

### 5.1 Patient Flow and Reliability of Trial

We will make a patient flow diagram as shown in Figure 1, following the CONSORT statement ^5^, to describe the progress of trial (enrollment, intervention, follow-up, and data analysis).

**Figure 1.**
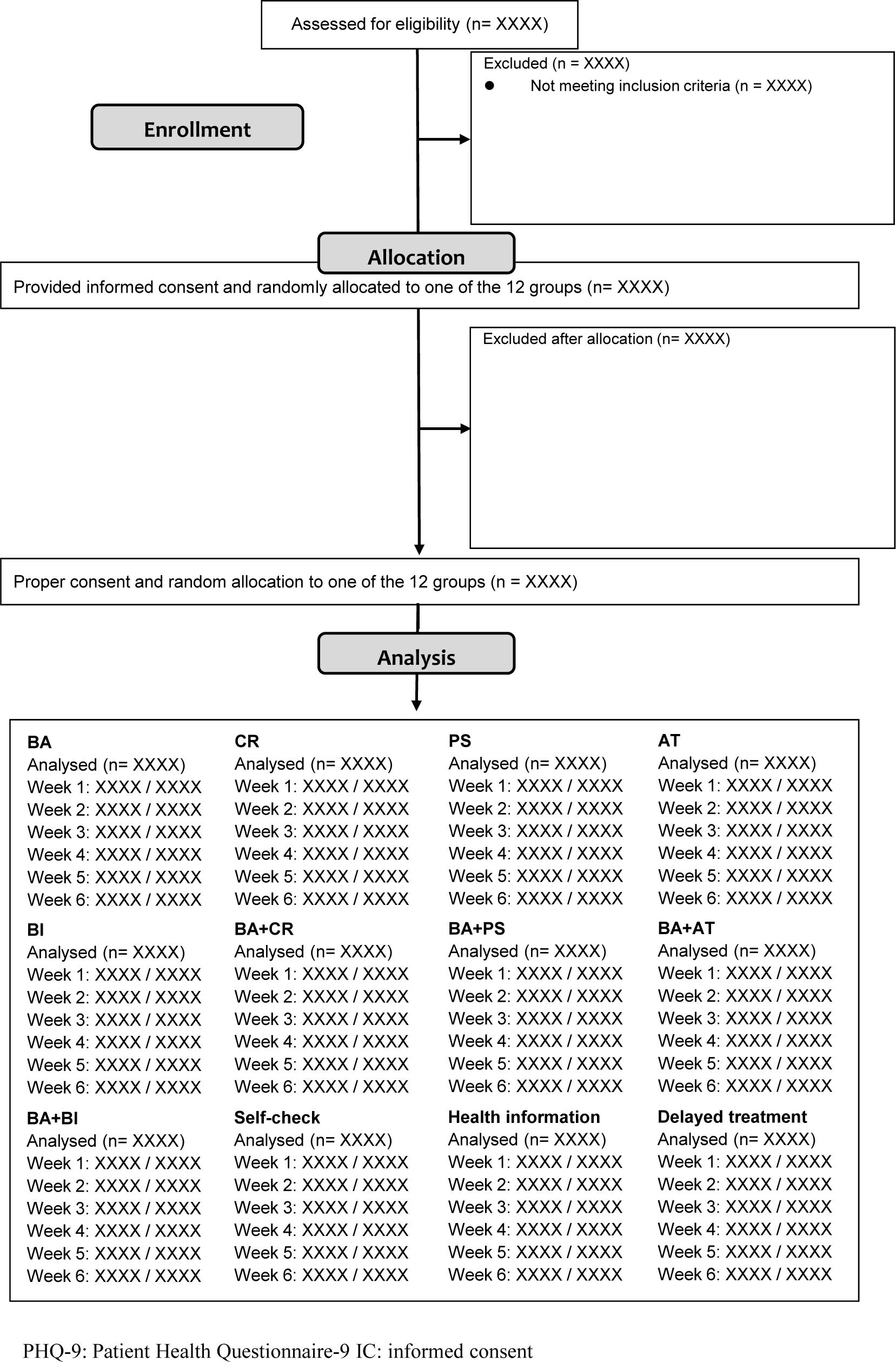
CONSORT diagram.

We will report the total number of patients assessed for eligibility, enrolled, and withdrawn/dropping out and their reasons, as well as frequency of inclusion/exclusion from the analysis and the reasons for exclusion.

### 5.2 Baseline Characteristics

We will summarize the participants’ baseline demographic, psychosocial and clinical characteristics in the ITT sample as shown in Table 3.

**Table 1.**
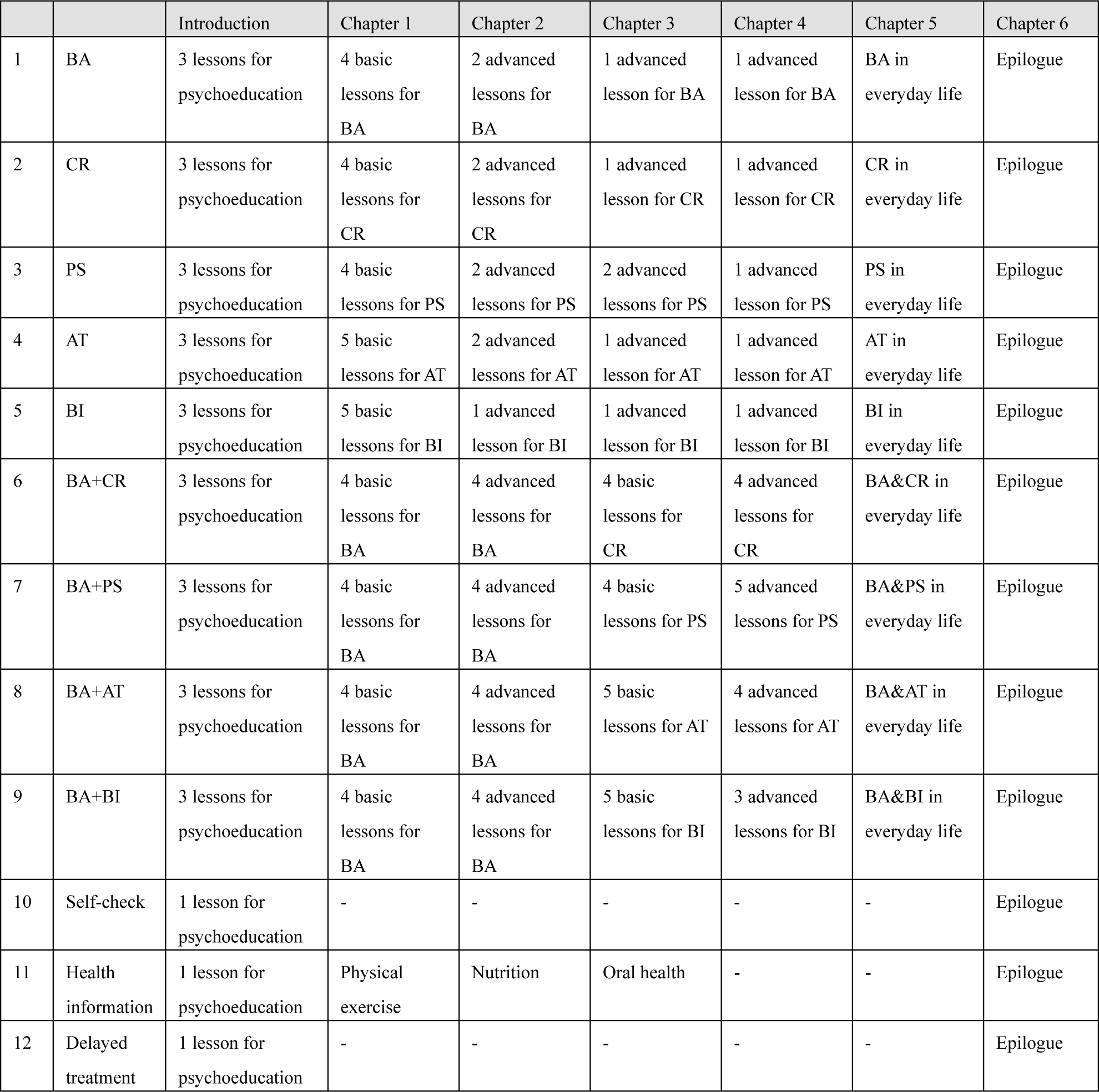
Contents of the interventions and controls.

**Table 2.** Measurement schedule.

|  | Questionnaire<br>Part I at landing<br>page | Questionnaire<br>Part II on<br>smartphone app<br>(Week 0) | Week 1 | Week 2 | Week 3 | Week 4 | Week 5 | Week 6 |
| --- | --- | --- | --- | --- | --- | --- | --- | --- |
| Allowances (days) | - |  | +6 | +6 | +6 | +6 | +6 | +14 |
| PHQ-9 | ○ | ○ | △ | △ | ○ | △ | △ | ○ |
| GAD-7 |  | ○ |  |  | ○ |  |  | ○ |
| ISI |  | ○ |  |  | ○ |  |  | ○ |
| SWEMWBS |  | ○ |  |  | ○ |  |  | ○ |
| CIDI |  | ○ |  |  |  |  |  |  |
| Baseline characteristics |  | ○ |  |  |  |  |  |  |
| ASC-Motivation |  | ○ |  |  | ○ |  |  |  |
| ICT usage |  | ○ |  |  |  |  |  |  |
| ACE |  | ○ |  |  |  |  |  |  |
| Short Big 5 |  | ○ |  |  |  |  |  |  |
| ASC-Life difficulties |  | ○ |  |  |  |  |  |  |
| ASC-Social support |  | ○ |  |  |  |  |  |  |
| CBT skills | ○ |  |  |  |  |  |  | ○ |
| WSAS |  | ○ |  |  |  |  |  | ○ |
| UWES |  | □ |  |  |  |  |  | □ |
| HPQ |  | □ |  |  |  |  |  | □ |
| EQ-5D-5L |  | ○ |  |  |  |  |  | ○ |
| Co-interventions |  |  |  |  |  |  |  | ○ |
| CSQ-3 |  |  |  |  |  |  |  | ○ |

○: to be administered to all participants, □: only to those currently employed, and △: all participants except for Groups 11 and 12.
ACE: Adverse Childhood Experiences, ASC: Assessment of Signal Cases, CIDI: Composite International Diagnostic Interview, CSQ-3: Customer Satisfaction Questionnaire-3, EQ-5D-5L: EuroQOL-5D-5L, GAD-7: General Anxiety Disorder-7, HPQ: Health and Work Performance Questionnaire, ICT: Information and communication technologies, ISI: Insomnia Severity Index, PHQ-9: Patient Health Questionnaire-9, SWEMWBS: Short Warwick Edinburgh Mental Well-Being Scale, UWES: Utrecht Work Engagement Scale, WSAS: Work and Social Adjustment Scale

**Table 3.**
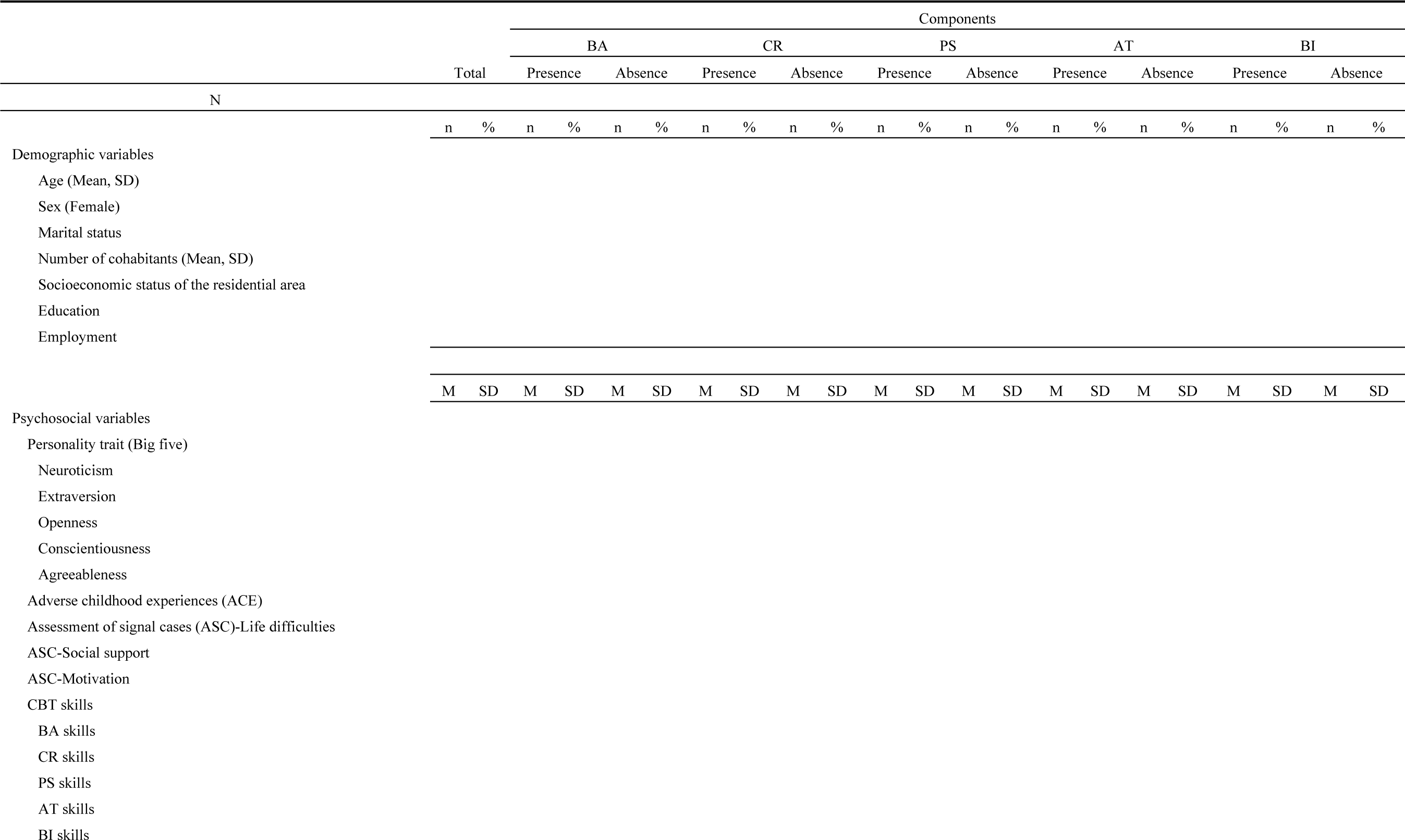
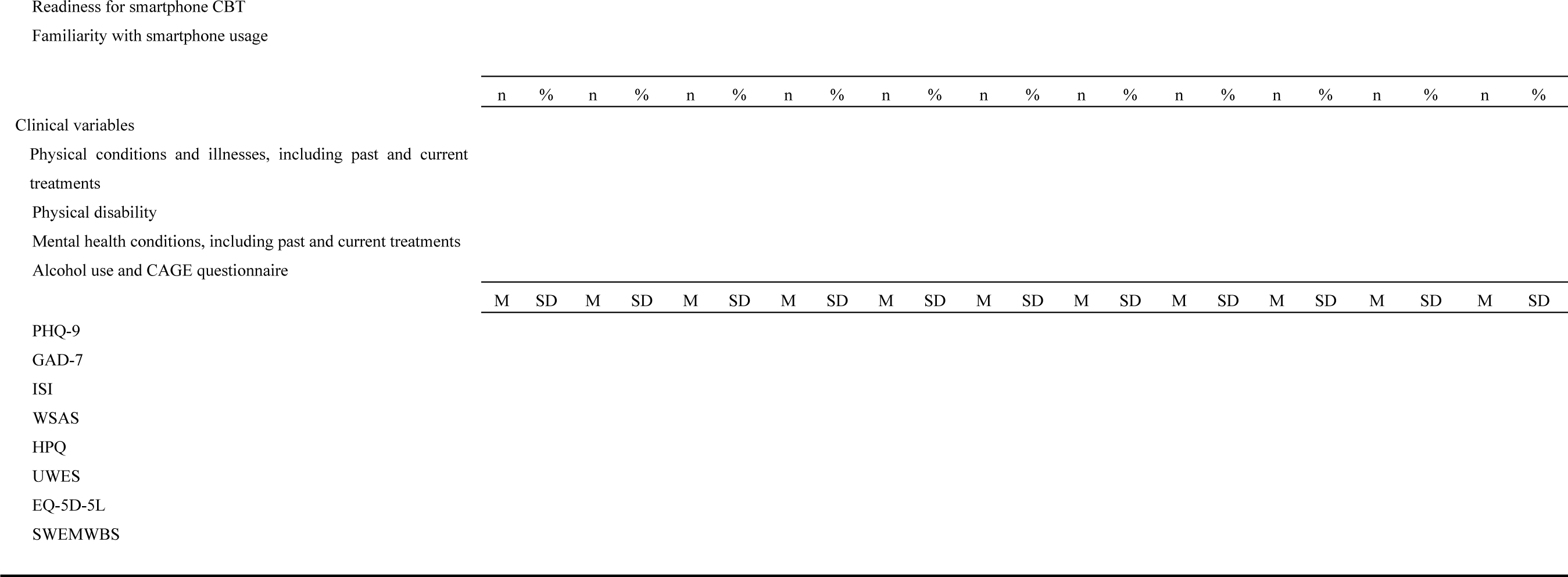
Baseline characteristics of total participants by each component (N= XXXX).

Continuous variables will be summarized using the number of observations (n), mean, standard deviation, median, minimum, and maximum. The number of missing observations will be reported.

For categorical variables, the frequencies and percentages (based on the non-missing sample size) as observed will be reported. The number of missing observations will also be reported.

## 6. Efficacy Analyses

### 6.1 Primary Efficacy Analyses

In the preplanned primary analysis, each of the four 2*2 factorial trials is to be analyzed independently. The four trials will use the BA arm (intervention arm #1 in Table 1) and the control arm (no treatment, intervention arm #12 in Table 1) in common. Intervention arms #10 and #11, with increasing stringency as control, will be used in sensitivity analyses, i.e., we will perform additional analyzes changing the control arm to the arm #10 or #11 respectively. For the fourth trial involving BA and BI, the participants who work in three shifts will be excluded, as shift workers cannot be expected to make optimal use of BI.

We will use the mixed-effects models for repeated-measures (MMRM) ^6,7^ to estimate the mean difference in change scores on the PHQ-9 for each component with adjusting informative missing of the outcomes. The model will include fixed effects of treatment, visit (as categorical), and treatment-by-visit interactions, adjusted for baseline PHQ-9 scores, employment status, age and sex. Each of the experimental factors will be coded at two levels (presence coded as 1 and absence coded as 0). The covariance matrix structure of outcome variables will be set to unstructured; if the estimating algorithm does not converge, we will use the matrix structure in the following order: Toeplitz, heterogeneous compound symmetry, autoregressive (1), compound symmetry, and variance components models. The standard error estimates and degree-of-freedom will be adjusted by the Kenward-Roger method ^8,9^. We will also estimate least squares mean change scores of the PHQ-9 at week 1, 2, 3, 4, and 5 based on the estimated models of the MMRM analysis. The results will be presented as Table 5. For the effect of BA, which will be estimated in all of the four 2*2 trials, we will present the effect size estimate of the first trial, i.e. the 2*2 trial involving BA and CR, in Table 5 as the primary result because BA and CR are the most typical ingredients across broadly conceived CBT interventions ^10^. However, if the effect size estimates of BA are considerably heterogeneous among the four trials, we will present all of these results, and will discuss the sources of the heterogeneity.

**Table 4.**
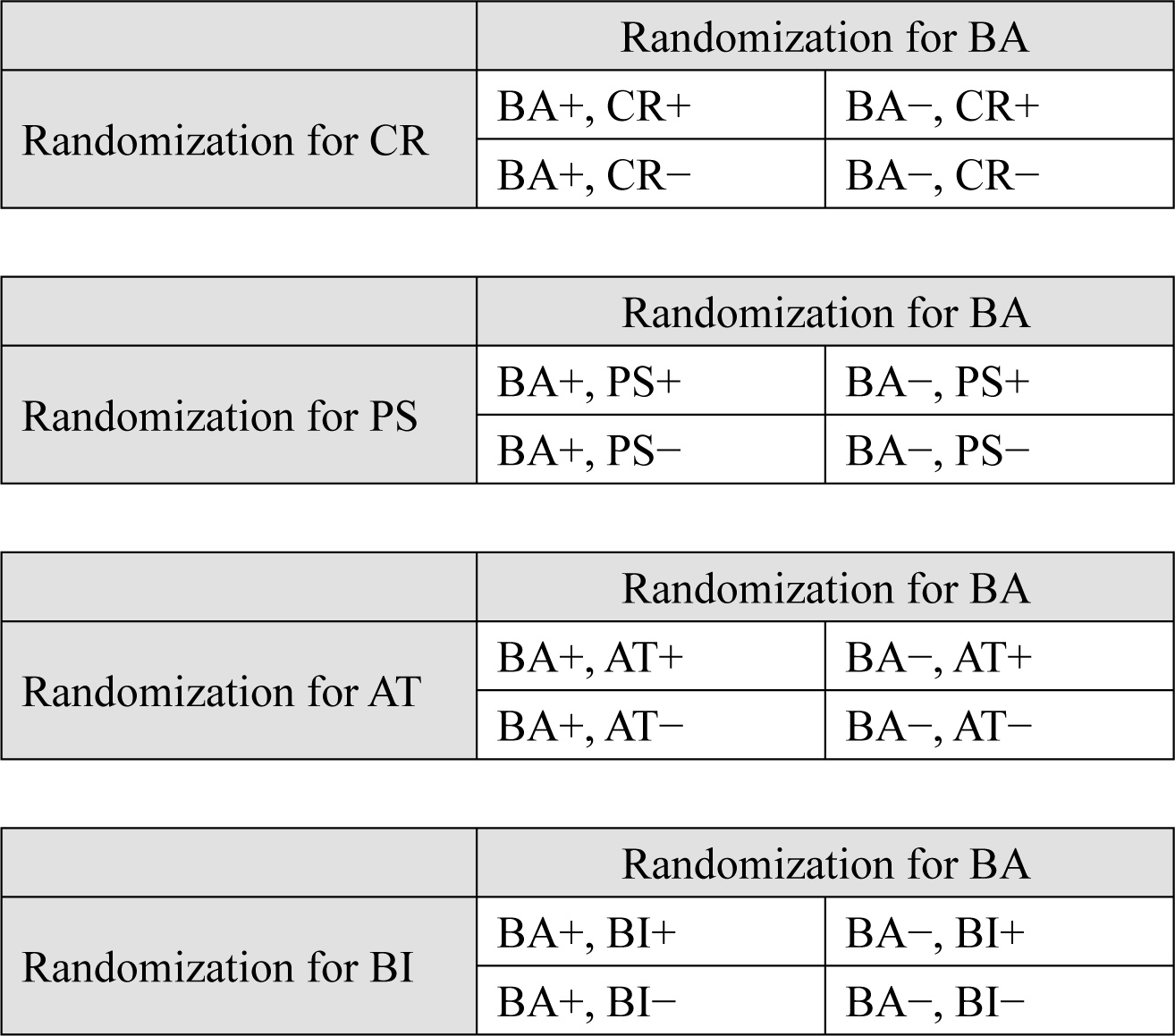
Four 2×2 factorial trials.

**Table 5.**
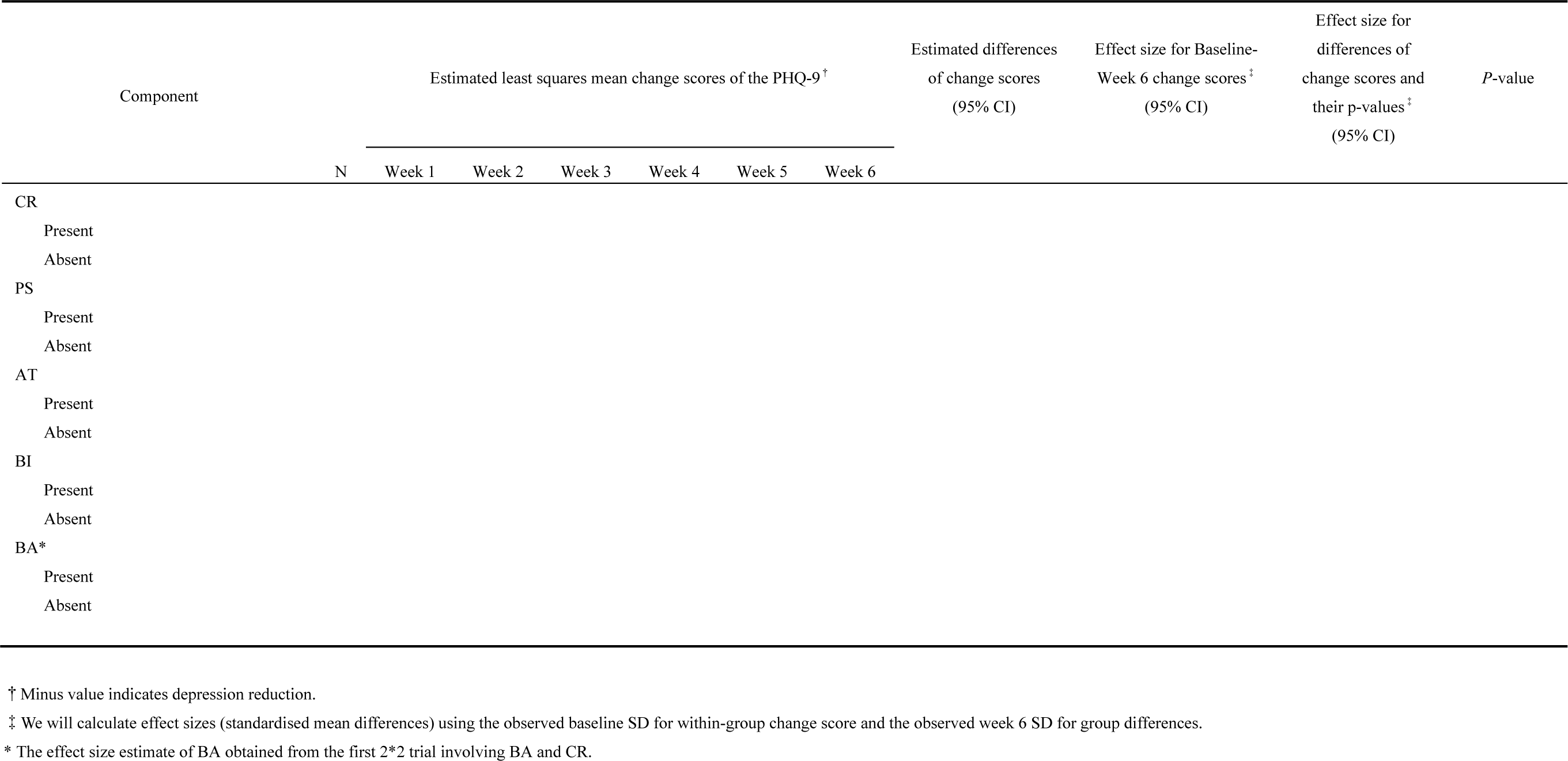
Analysis of main effects using the mixed-model repeated measures analysis (MMRM) for PHQ-9 score.

No adjustment for multiple testing will be applied in examination of statistical significance of the main effects and the conventional threshold for statistical significance (p<0.05, two sided) will be used, because conventionally, the multiple hypotheses have been tested independently in these trial designs assuming as if the tested interventions could have been assessed in separate trials ^11,12^. We will perform another MMRM analysis involving a two-way interaction term of each two components to assess the interaction between the treatments as a sensitivity analysis (Table 6). Previous research has not been suggestive of interactions among iCBT components ^10,13^, but when we identify a strong interaction, we will interpret the results considering the interaction. We also perform linear regression analyses using only the week 6 data as sensitivity analyzes.

**Table 6.**
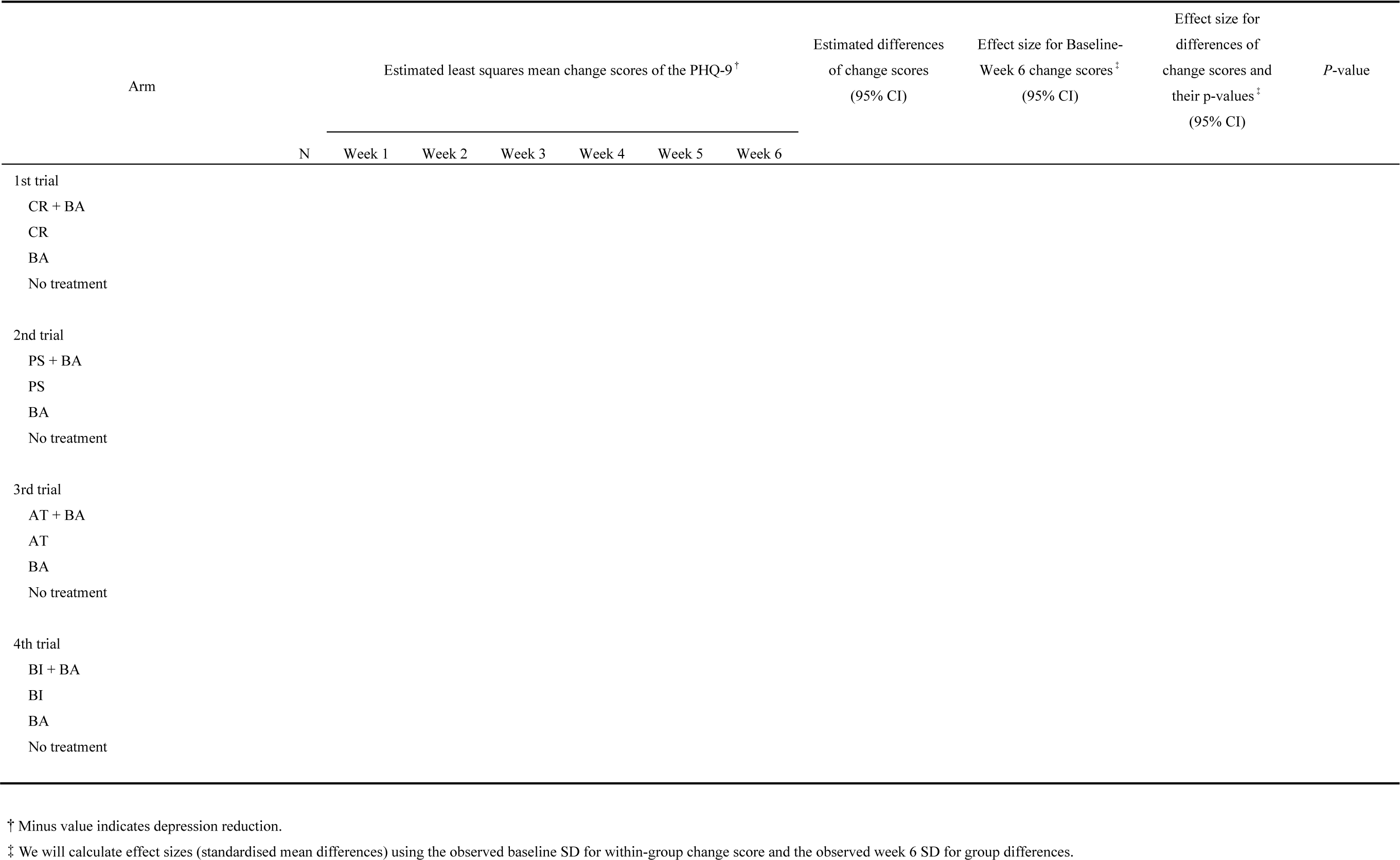
Analysis of main and interaction effects using the mixed-model repeated measures analysis (MMRM) for PHQ-9 score.

### 6.2 Secondary Efficacy Analyses

#### 6.1.1 Combined Master Protocol Analysis via Pooling the Four Factorial Trials

To assess the comparative efficacy of the five CBT skills and their combinations, we will additionally perform a combined master protocol analysis via pooling the four factorial trials along with all the control arms. We will perform the MMRM analysis ^6,7^ for the pooled dataset, and will provide comparative effect size information for all the CBT skills and their combinations evaluated in the four trials. We will perform two combined master protocol analyses involving (1) main effects of 5 CBT skills and (2) main effects and 2 way interaction terms of 5 CBT skills, while setting the control arm to the arm #12. Control arms #10 and #11, whose outcomes are assumed to be different, will also be included and compared comprehensively. The results will be presented as in Table 7. The combined master protocol analysis allows direct comparisons among all active components, thus informing (1) what component is the most efficacious and (2) whether the effects of individual CBT skills are different (via significance tests). In this combined master protocol analysis, we will include shift-workers, although such inclusion may lead to underestimating the effect of BI on depression, to maintain the statistical power for the analysis and make the results as comparable as possible to those in the primary 2*2 trial analyses.

**Table 7.**
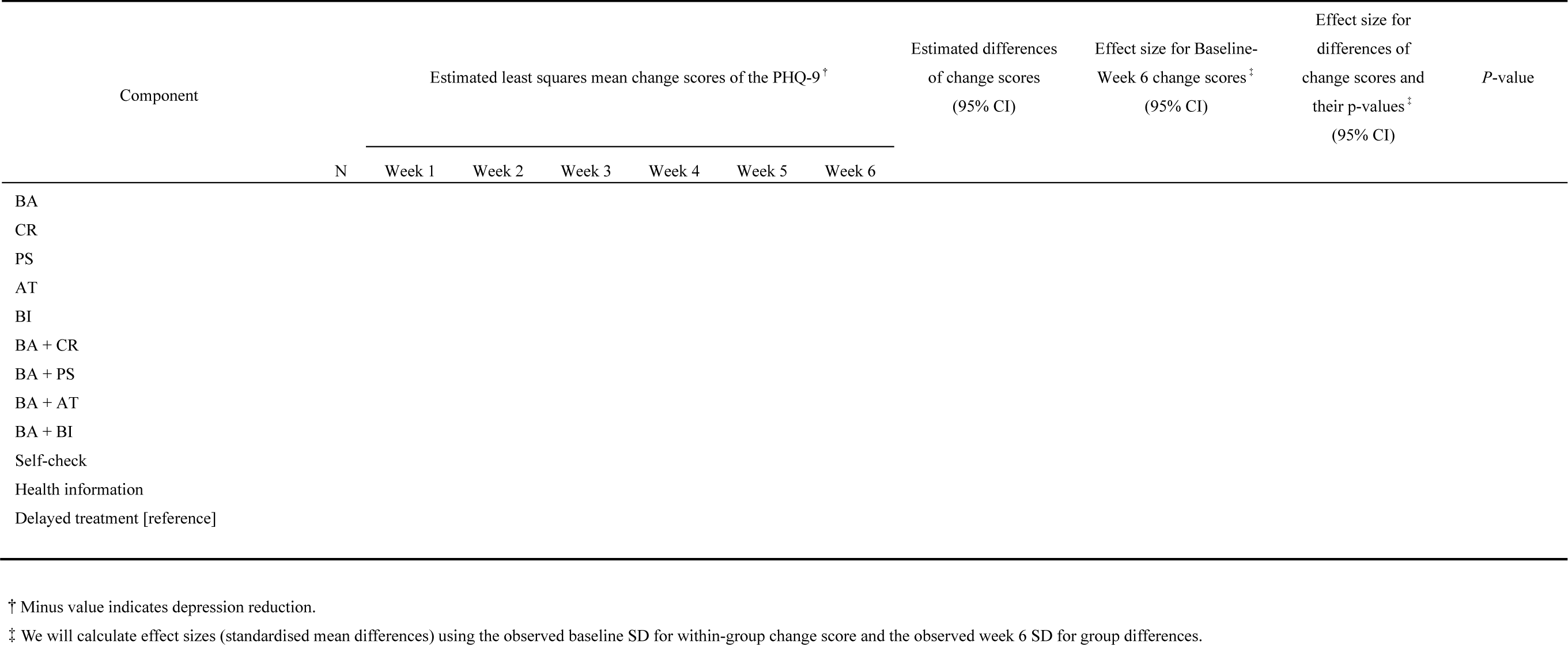
Combined master protocol analysis of main and interaction effects using the mixed-model repeated measures analysis (MMRM) for PHQ-9 score.

#### 6.2.2 Efficacy Analyses for Secondary Outcomes

We will perform MMRM analyzes for the repeated measurement outcomes of GAD-7, ISI and SWEMWBS until week 6. The same modelling strategy with the analysis of the primary outcome in Section 6.1 will be adopted.

#### 6.2.3 Sensitivity analyses

We will evaluate the robustness of the foregoing analyses by conducting the following sensitivity analyses.

For the 2*2 trials

(1) Using the control arm #10 (self-check) or #11 (health information) instead of #12 (delayed treatment) in each of the 2*2 trial
(2) We will examine interactions among the skills as a sensitivity analysis. In the examination of interactions among components, we will also perform linear regression analyses using only the week 6 data for each of the 2*2 trials.
(3) For the factorial trial examining BA and BI, subgrouping participants to those who scored 8 or more vs those who scored 7 or less on the ISI at baseline. Here the primary outcome is the PHQ-9, and this subgroup analysis will confirm if the effect of BI is similar regardless of the baseline insomnia severity.

For the secondary outcomes

(4) For the secondary outcome of the GAD-7, limiting participants to those who scored 5 or more on the GAD-7 at baseline (i.e. those who presented with at least some anxiety symptoms at baseline). This will mitigate the floor effect on the GAD-7.
(5) For the secondary outcome of the ISI, limiting participants to those who scored 8 or more on the ISI at baseline (i.e. those who presented with at least some insomnia symptoms at baseline). This will mitigate the floor effect on the ISI.
(6) For the secondary outcome of the SWEMWEBS in the four 2*2 factorial trials, limiting participants to those who scored 20 or less on the SWEMWBS at baseline (i.e. those who showed reduced well-being at baseline). This will mitigate the ceiling effect on the SWEMWEBS.

## 7. Safety Analyses

As this trial is a non-invasive interventional study and no adverse events due to the intervention are expected. We will only descriptively report serious adverse events as reported during the 6-week intervention in each arm.

## 8. Revision History

| Version | Date | Changes |
| --- | --- | --- |
| 1.1 | 03-16-2024 | The first version. |

## Data Availability

This is a statistical analysis plan document and does not involve new data.

## ABBREVIATIONS AND DEFINITIONS

CBT: cognitive-behavioral therapy
CR: cognitive restructuring
BA: behavioural activation
PS: problem-solving
AT: assertion training
BI: behavior therapy for insomnia
PHQ-9: Patient Health Questionnaire-9
GAD-7: Generalized Anxiety Disorder-7
ISI: Insomnia Severity Index
SWEMWBS: Short Warwick Edinburgh Mental Well-Being Scale
WSAS: Work and Social Adjustment Scale
UWES: Utrecht Work Engagement Scale
HPQ: Health and Work Performance Questionnaire
EQ-5D-5L: EuroQOL-5D-5L
CAQ-3: Client Satisfaction Questionnaire-3
FAS: full analysis set
MMRM: mixed-effects models for repeated measures

